# Post-COVID-19 syndrome in outpatients: a cohort study

**DOI:** 10.1101/2021.04.19.21255742

**Authors:** Florian Desgranges, Eliana Tadini, Aline Munting, Jean Regina, Paraskevas Filippidis, Benjamin Viala, Eleftherios Karachalias, Véronique Suttels, David Haefliger, Eleftheria Kampouri, Mathias Van Singer, Jonathan Tschopp, Laurence Rochat Stettler, Siméon Schaad, Thomas Brahier, Olivier Hugli, Yolanda Mueller Chabloz, Alexandre Gouveia, Onya Opota, Pierre-Nicolas Carron, Benoît Guery, Matthaios Papadimitriou-Olivgeris, Noémie Boillat-Blanco, the RegCOVID research group

**Author notes:** Equal contribution to this work (Matthaios Papadimitriou-Olivgeris, Noémie Boillat-Blanco). Members of the RegCOVID group are listed in the acknowledgement section. **Corresponding author** Dr. Florian Desgranges, Service des maladies infectieuses, Centre hospitalier universitaire vaudois (CHUV), Rue du Bugnon 46, CH-1011 Lausanne, Switzerland,; 0041795561721.

## Abstract

**Background:** After mild COVID-19, some outpatients experience persistent symptoms. However, data are scarce and prospective studies are urgently needed.

**Objectives:** To characterize the post-COVID-19 syndrome after mild COVID-19 and identify predictors.

**Participants:** Outpatients with symptoms suggestive of COVID-19 with (1) PCR-confirmed COVID-19 (COVID-positive) or (2) SARS-CoV-2 negative PCR (COVID-negative).

**Design:** Monocentric cohort study with prospective phone interview between more than three months to ten months after initial visit to the emergency department and outpatient clinics.

**Main Measures:** Data of the initial visits were extracted from the electronic medical file. Predefined persistent symptoms were assessed through a structured phone interview. Associations between long-term symptoms and PCR results, as well as predictors of persistent symptoms among COVID-positive, were evaluated by multivariate logistic regression adjusted for age, gender, smoking, comorbidities, and timing of the survey.

**Key results:** The study population consisted of 418 COVID-positive and 89 COVID-negative patients, mostly young adults (median age of 41 versus 36 years in COVID-positive and COVID-negative, respectively; *p=*0.020) and health care workers (67% versus 82%; *p=*0.006). Median time between the initial visit and the phone survey was 150 days in COVID-positive and 242 days in COVID-negative patients. Persistent symptoms were reported by 223 (53%) COVID-positive and 33 (37%) COVID-negative patients (*p=*0.006). Overall, 21% COVID-positive and 15% COVID-negative patients (*p=*0.182) attended care for this purpose. Four surveyed symptoms were independently associated with COVID-19: fatigue (adjusted odds ratio [or] 2.14, 95%CI 1.04-4.41), smell/taste disorder (26.5, 3.46-202), dyspnea (2.81, 1.10-7.16) and memory impairment (5.71, 1.53-21.3). Among COVID-positive, female gender (1.67, 1.09-2.56) and overweight/obesity (1.67, 1.10-2.56) were predictors of persistent symptoms.

**Conclusions:** More than half of COVID-positive outpatients report persistent symptoms up to ten months after a mild disease. Only 4 of 14 symptoms were associated with COVID-19 status. The symptoms and predictors of the post-COVID-19 syndrome need further characterization as this condition places a significant burden on society.

## INTRODUCTION

Almost two years after the first cases of Coronavirus disease 2019 (COVID-19) (1), the severe acute respiratory syndrome coronavirus 2 (SARS-CoV-2) pandemic remains a constant challenge for healthcare systems and affects the well-being of many individuals. The clinical presentation and outcomes of acute COVID-19 are well described. Most patients have mild disease and only a minority need hospital admission (2). In most cases, patients experience a complete resolution of their symptoms after two to six weeks (3), but a subgroup present long lasting symptoms. These persistent symptoms have been described in observational studies (4,5) and cohorts with a large proportion of outpatients (6,7). Three studies reported that an important number of patients had persistent symptoms for more than six months after COVID-19: one study included adults discharged from hospital (8) and two studies focused on outpatients with mild-to-moderate disease (9,10). NICE guideline currently defines post-COVID-19 syndrome as the persistence of signs and symptoms for more than three months after infection, in the absence of an alternative diagnosis (11).

To the best of our knowledge, no study evaluated the prevalence of persistent symptoms among patients with symptoms suggestive of COVID-19, comparing patients positive for SARS-CoV-2 to those tested negative. Furthermore, no study explored the factors associated with persistent symptoms in COVID-19 outpatients. These data may modify patient’s management and influence public health vaccination strategies by identifying additional population groups that should be prioritized.

We aimed to compare the prevalence of symptoms persistent for more than three months between SARS-CoV-2 PCR-confirmed (COVID-positive) and PCR-negative (COVID-negative) patients in outpatient clinics, and to identify predictors of persistent symptoms in COVID-positive.

## METHODS

### Study design, setting and participants

This cohort study recruited consecutive PCR-confirmed COVID-19 patients (COVID-positive) during the initial visit in the emergency department (ED), in the SARS-CoV-2 screening center and in two outpatient clinics of the University of Lausanne, Switzerland, between February 26^th^ and April 27^th^, 2020. During the study period, SARS-CoV-2 testing was restricted to patients with symptoms suggestive of COVID-19 (i.e., symptoms of acute respiratory tract infection, history of fever or sudden loss of smell or taste) (1) with at least one risk factor for severe COVID-19 (i.e., ≥ 65 years old or presence of a comorbidity such as hypertension, diabetes, cancer, chronic cardiac or respiratory disease, immunosuppression) or (2) working as health care workers (HCW). Validated nucleic acid amplification tests were used and a dedicated trained medical team performed nasopharyngeal swabs (12).

We included a control group of outpatients with a negative nasopharyngeal swab PCR SARS-CoV-2 result (COVID-negative). This group was recruited in the same health facilities during the same period with the same screening criteria (≥ 1 risk factor for severe COVID-19 or HCW) as COVID-positive. The only differences between the two groups were the types of symptoms at inclusion. The clinical inclusion criteria of COVID-negative were the presence of cough, dyspnea, or history of fever while criteria of COVID-positive were the presence of any symptom suggestive of COVID-19. This difference is related to the fact that we used the COVID-negative group of a previously described prospective cohort (13,14). The use of this pre-existing cohort also influenced the number of COVID-negative patients included.

We excluded patients who 1) did not have a documented health assessment at the time of the test, 2) were hospitalized for more than 24 hours on the day of initial visit or within the following 30 days, 3) were unable to understand the phone survey due to a language barrier or a cognitive impairment, 4) declined to participate, 5) were not reachable for the phone interview after at least two attempts, or 6) had the phone survey less than 3 months after the initial visit by mistake. Patients with a positive SARS-CoV-2 test performed in another location before the initial visit in the study centers were excluded, as we also planned to evaluate the impact of the infection on the healthcare system (i.e., medical consultations for persistent symptoms). Moreover, we excluded COVID-negative patients who declared having a documented (PCR or serology) SARS-CoV-2 infection before the phone follow-up.

### Procedures

We retrospectively extracted data from the patients’ electronic medical records: demographics, medical history, initial clinical presentation, and SARS-CoV-2 real-time reverse transcriptase PCR (RT-PCR) cycle thresholds. We prospectively conducted a structured and standardized phone survey more than three months after the initial consultation. We collected information on 1) persistent symptoms at the time of the call, 2) secondary hospital admission within 30 days of the initial consultation, 3) medical consultation for persistent symptoms until the call, and 4) anthropometric data. The survey included 14 pre-defined symptoms: fatigue, muscle weakness, dyspnea, cough, thoracic pain, smell or taste disorder, blurred vision, headache, memory impairment, loss of balance, numbness, nausea, sleep disorder, and hair loss. Dyspnea was further categorized according to New York Heart Association (NYHA) grades (15).

Physicians of the infectious diseases service conducted the phone survey. As they did not have dedicated time to perform these calls, they did not survey all patients at the same time point after the initial consultation. Clinicians called COVID-positive patients randomly. They called COVID-positive patients first. Indeed, we decided to include a group of COVID-negative patients in a second phase when we realized that a large proportion of COVID-positive participants had persistent symptoms. To account for the timing of symptoms assessment, we classified COVID-positive patients in three equivalent 2-month periods according to the time between SARS-CoV-2 diagnosis (initial consultation) and the phone interview: first period: >3 months to 5 months after diagnosis, second period: >5 months to 7 months after diagnosis, and third period: >7 months to 10 months after diagnosis.

All data were entered in the Lausanne University Hospital’s electronic database “regCOVID” using the REDCap® platform (Research Electronic Data Capture v8.5.24, Vanderbilt University, Tennessee, USA) (16).

### Definitions

Long-term symptoms were defined as symptoms that appeared or worsened during the acute infection phase, were still present at time of the phone call (i.e., more than three months after the initial consultation) and were not explained by an alternative diagnosis. Smell or taste disorder included partial or complete loss of that sense. Numbness included hypoesthesia and paresthesia.

### Statistical analyses

Demographics, clinical features at the time of initial consultation, consultation for persistent symptoms, and prevalence of long-term symptoms (any symptom or individualized symptoms) were compared between COVID-positive and COVID-negative by Mann Whitney-U, *chi*-squared, or Fisher tests as appropriate, as well as between COVID-positive patients surveyed during the three different periods after SARS-CoV-2 diagnosis by 1-way ANOVA, Kruskal-Wallis, or Chi-squared as appropriate. A *p* value <0.05 was considered statistically significant. The association between COVID-19 and long-term symptoms was evaluated by multivariate logistic regression including potential confounders (age, gender, smoking habits, comorbidities, period of the phone survey). To account for the difference in the timing of symptoms assessment between COVID-positive and COVID-negative, we added a subgroup analysis including COVID-positive and COVID-negative patients surveyed during the same period (the third period). The independent association between the previously mentioned factors and each long-term symptom in COVID-positive was evaluated by multivariate logistic regression including the same potential confounders. To account for the large number of HCW in the study population, we added a subgroup analysis including only HCW. STATA (version 15.1, Stata Corp, College Station, TX, USA) statistical software and GraphPad Prism 8.3 (GraphPad, San Diego, CA, USA) were used for analyses.

### Ethics

The project was approved by the Ethics Committee of the Canton of Vaud, Switzerland. All participants gave their verbal consent to participate in this study during the phone interview (project-ID CER-VD 2020-01107 and 2019-02283).

## RESULTS

A total of 956 symptomatic patients had a RT-PCR-confirmed COVID-19 between February 26^th^ and April 27^th^, 2020 and 538 patients were excluded (Figure 1). The remaining 418 (44%) COVID-positive patients were included in the analysis. 131 symptomatic COVID-negative patients with a negative SARS-CoV-2 RT-PCR between March 31^st^ and April 26^th^, 2020 were included. 19 patients with a positive SARS-CoV-2 test (RT-PCR or serology) between the initial visit and the phone survey, 17 who could not be reached, 3 who refused to participate, 2 who did not speak one of the investigators’ languages and 1 who died were excluded. The remaining 89 (68%) COVID-negative patients were included in the analysis.

**Figure 1:**
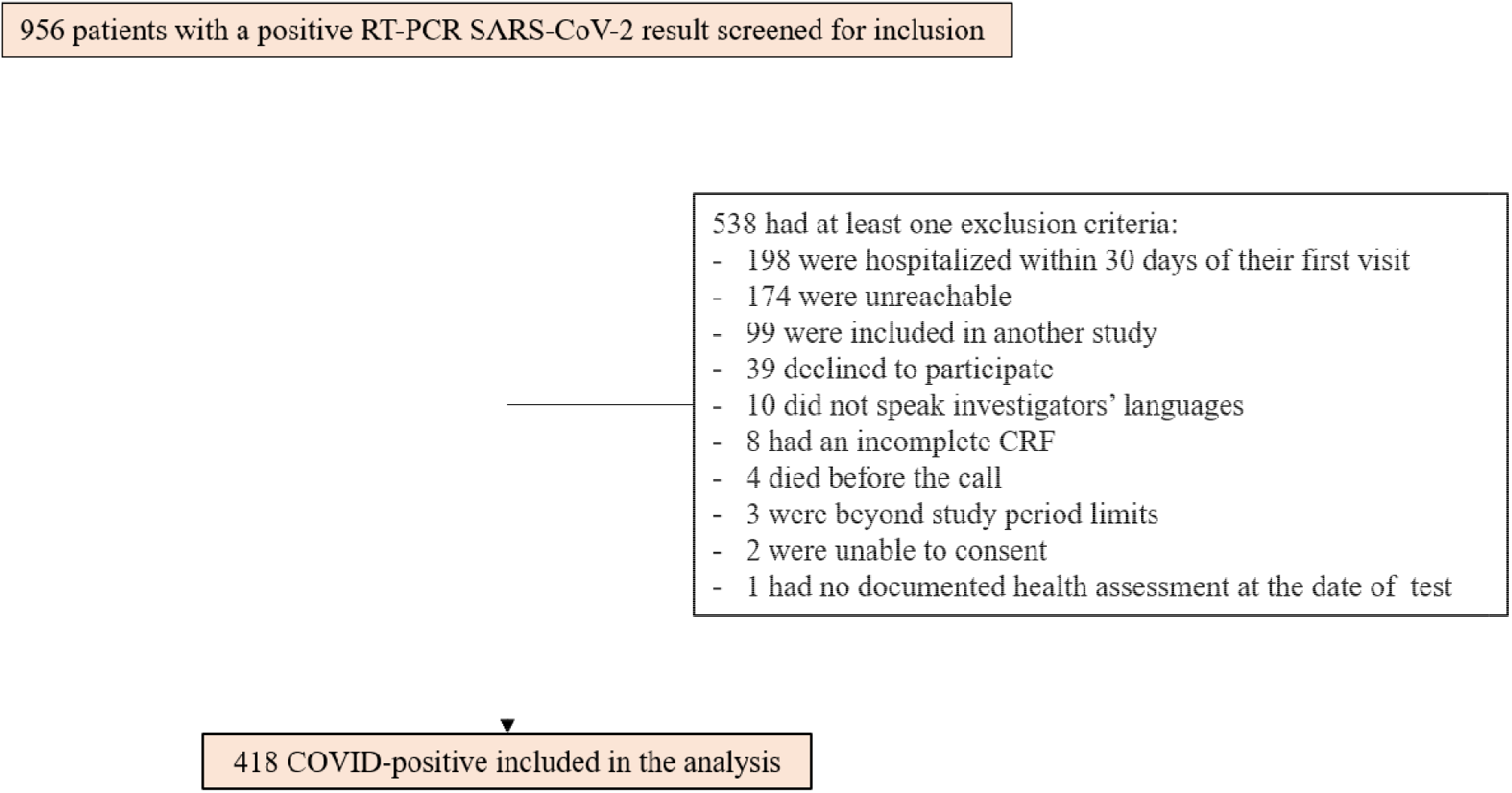
Study flow-chart.

### Demographics and clinical presentation

In both groups, there was a majority of women and HCW. Approximately a third of patients had a comorbidity, mainly obesity, arterial hypertension, and asthma (Table 1). Most patients were tested within the first week of symptom onset and most had normal vital signs at initial consultation. Compared to COVID-negative, COVID-positive were slightly older and less often HCW, active smokers, or asthmatic. COVID-positive reported less often a history of fever, dyspnea, or sore throat at the time of evaluation.

**Table 1:**
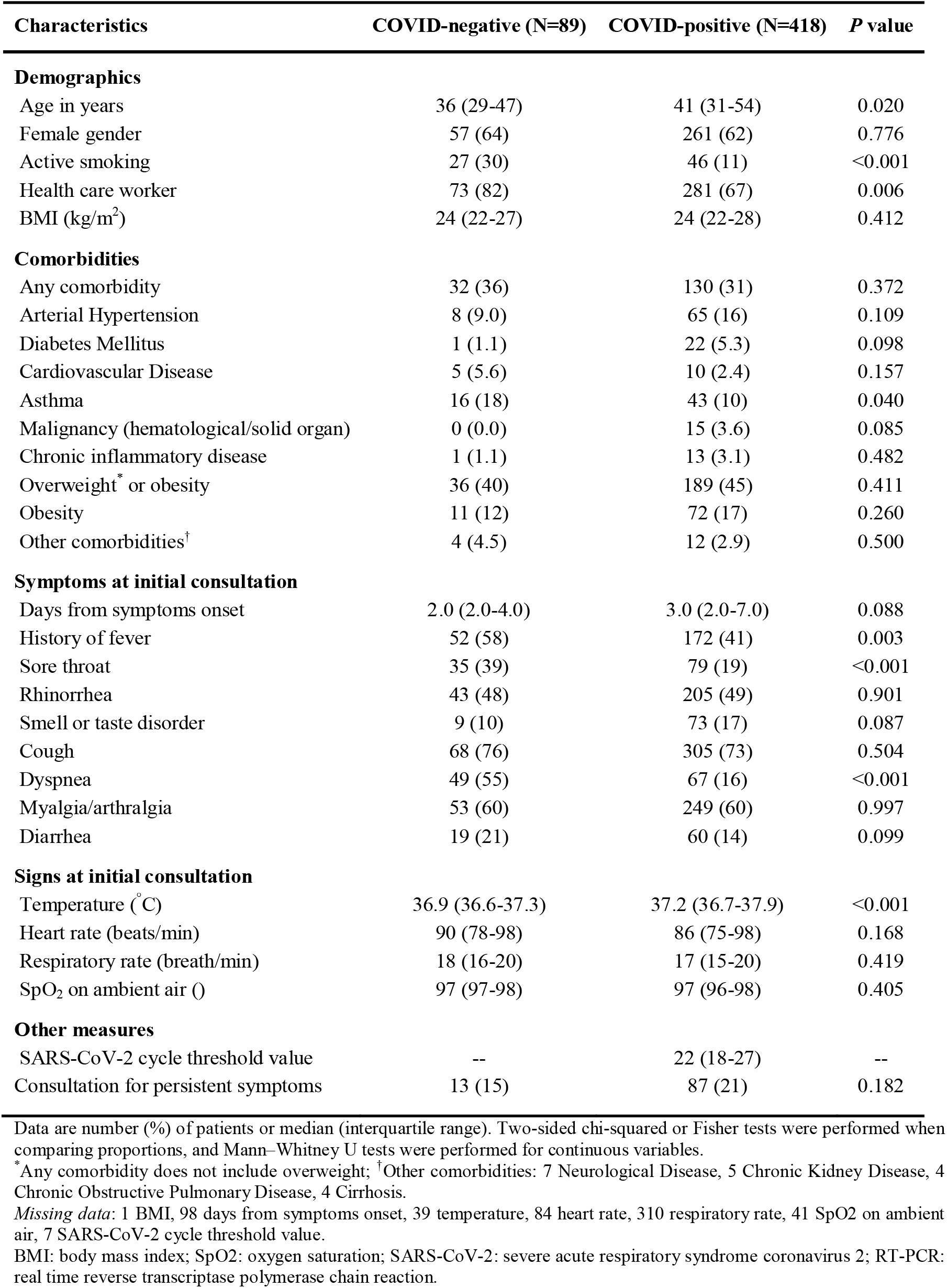
Characteristics of patients according to SARS-CoV-2 RT-PCR result (COVID-negative and COVID-positive).

The study team assessed long-term symptoms via the phone survey at a median of 150 days (interquartile range [IQR], 121-204 days) and 242 days (IQR 236-248 days) after the initial consultation in the COVID-positive and COVID-negative groups, respectively (*p*<0.001). The study team called 190 (45%) COVID-positive during the first period (>3 to 5 months after the initial visit), 102 (24%) during the second period (>5 to 7 months) and 126 (30%) during the third period (>7 to 10 months). Patients called in the first period were significantly older, less often HCW, and had more comorbidities than those surveyed during the two other periods (Appendix table 1). The study team called all COVID-negative during the third period.

### Persistent symptoms

At the time of the phone survey, 223/418 (53%) COVID-positive and 33/89 (37%) COVID-negative patients (*p=*0.006) reported the presence of any long-term symptom (Figure 2A, Table 2).

**Table 2.** Association between long-term symptoms (any and individualized symptoms) assessed at the time of phone survey and COVID positive status.

| Long-term symptoms | COVID-negative<br>(n=89) | COVID-positive<br>(n=418) | Bivariate analysis* |  | Multivariate analysis*<sup>†</sup> |  |
| --- | --- | --- | --- | --- | --- | --- |
|  |  |  | OR<br>(95% CI) | P value | aOR<br>(95% CI) | P value |
| Any symptoms | 33 (37) | 223 (53) | 1.94<br>(1.21-3.11) | 0.006 | 2.04<br>(1.14-3.67) | 0.017 |
| Fatigue | 15 (17) | 132 (32) | 2.28<br>(1.26-4.12) | 0.006 | 2.14<br>(1.04-4.41) | 0.039 |
| Muscle weakness | 2 (2.2) | 8 (1.9) | 0.85<br>(0.18-4.07) | 0.837 | 0.65<br>(0.08-5.21) | 0.688 |
| Dyspnea | 7 (7.9) | 66 (16) | 2.20<br>(0.97-4.96) | 0.059 | 2.81<br>(1.10-7.16) | 0.030 |
| Dyspnea NYHA≥2 | 4 (4.5) | 30 (7.2) | 1.64<br>(0.56-4.79) | 0.363 | 1.11<br>(0.29-4.32) | 0.881 |
| Cough | 3 (3.4) | 25 (6.0) | 1.82<br>(0.54-6.2) | 0.334 | 1.16<br>(0.24-5.56) | 0.849 |
| Thoracic pain | 4 (4.5) | 21 (5.0) | 1.12<br>(0.38-3.36) | 0.834 | 1.11<br>(0.27-4.54) | 0.886 |
| Smell or taste disorder | 1 (1.1) | 93 (22) | 25.2<br>(3.46-183) | 0.001 | 26.5<br>(3.46-202) | 0.002 |
| Blurred vision | 2 (2.2) | 20 (4.8) | 2.19<br>(0.50-9.53) | 0.298 | 3.17<br>(0.63-16.0) | 0.163 |
| Headache | 10 (11) | 50 (12) | 1.07<br>(0.52-2.21) | 0.847 | 1.27<br>(0.52-3.09) | 0.601 |
| Loss of balance | 3 (3.4) | 12 (2.9) | 0.85<br>(0.23-3.07) | 0.801 | 0.37<br>(0.05-2.62) | 0.322 |
| Memory impairment | 3 (3.4) | 48 (11) | 3.72<br>(1.13-12.2) | 0.003 | 5.71<br>(1.53-21.3) | 0.010 |
| Numbness | 1 (1.1) | 14 (3.4) | 3.05<br>(0.40-23.5) | 0.284 | 1.71<br>(0.14-20.3) | 0.670 |
| Nausea | 3 (3.4) | 11 (2.6) | 0.77<br>(0.21-2.84) | 0.700 | 1.30<br>(0.24-7.08) | 0.760 |
| Sleep disorder | 10 (11) | 41 (9.8) | 0.86<br>(0.41-1.79) | 0.685 | 1.03<br>(0.42-2.52) | 0.950 |
| Hair loss | 5 (5.6) | 43 (10) | 1.92<br>(0.74-5.01) | 0.179 | 1.18<br>(0.36-3.93) | 0.783 |

Data are number (%) of patients.
\*symptoms associated with COVID-positive status. <sup>†</sup>Analysis adjusted for age, gender, smoking, overweight/obesity, diabetes, asthma, hypertension, cancer, cardiovascular disease, chronic inflammatory disease, timing of the phone survey.
OR: odds ratio; aOR: adjusted odds ratio; CI: confidence interval; NYHA: New York Heart Association.

**Figure 2:**
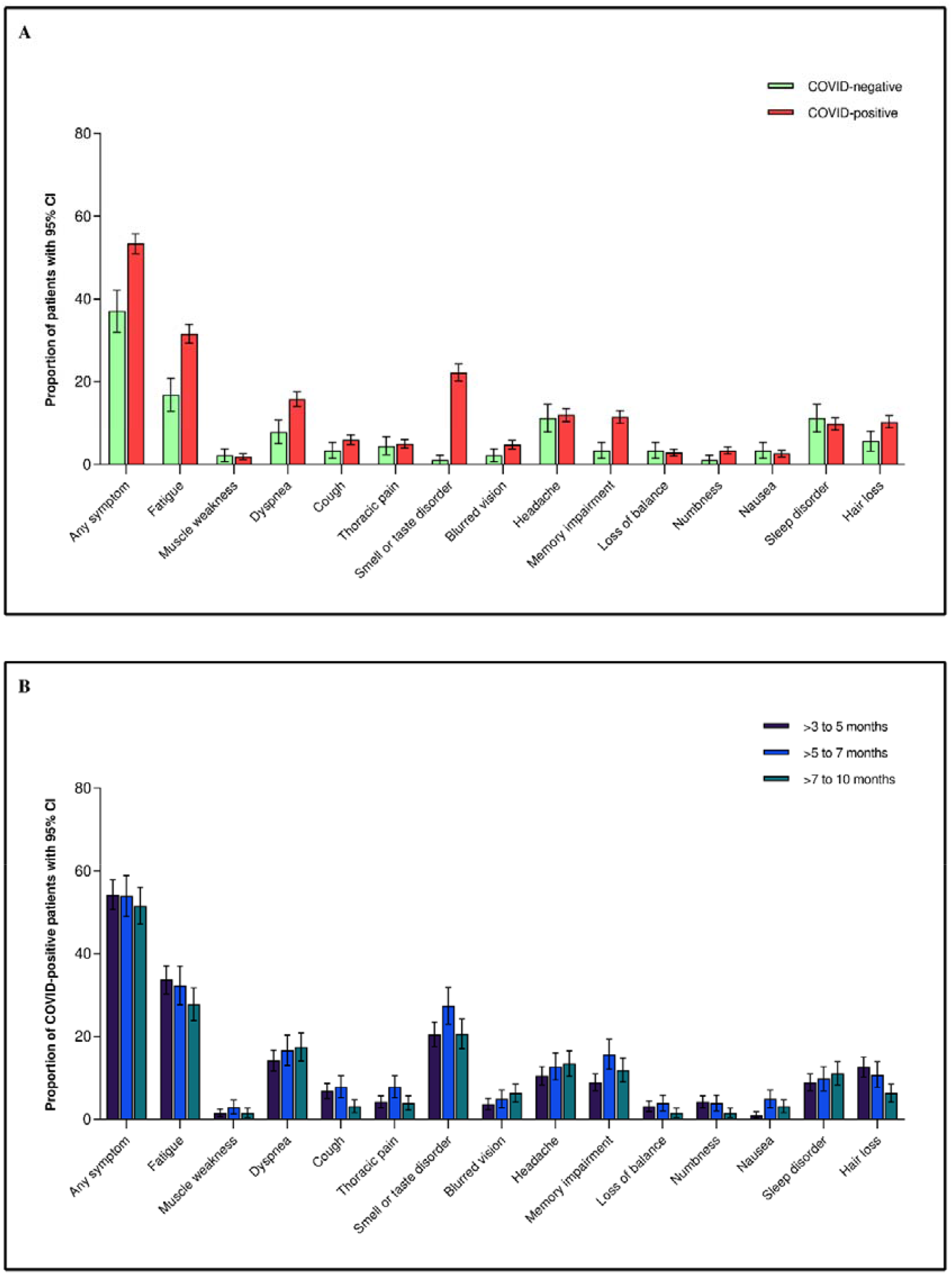
Proportions of patients with long-term symptoms at the time of the phone survey. **A)** Comparison between COVID-positive (positive SARS-CoV-2 RT-PCR) and COVID-negative (negative SARS-CoV-2 RT-PCR); **B)** Comparison between COVID-positive surveyed during three different period: Period 1: >3 to 5 months after SARS-CoV-2 diagnosis; Period 2: >5 to 7 months after SARS-CoV-2 diagnosis; Period 3: >7 to 10 months after SARS-CoV-2 diagnosis. Data are shown as proportions (%) with 95% confidence intervals.

In COVID-positive, the main reported symptoms were fatigue (n=132, 32%), smell or taste disorder (n=93, 22%), dyspnea (n=66, 16%), headache (n=50, 12%), memory impairment (n=48, 11%), hair loss (n=43, 10%), and sleep disorders (n=41, 10%), irrespective of HCW status (Appendix table 3B). The prevalence of long-term symptoms was similar during the three time periods (Figure 2B). In COVID-negative, the main reported symptoms were fatigue (n=15, 17%), headache (n=10, 11%), sleep disorders (n=10, 11%), and dyspnea (n=7, 8%). COVID-positive reported more often smell or taste disorder (*p=*0.001), fatigue (*p=*0.006), and memory impairment (*p=*0.003) compared to COVID-negative, irrespective of the timing of the survey (Appendix table 2). The prevalence of other symptoms was similar between COVID-positive and COVID-negative. The proportions of patients with ≥ 2 and ≥ 3 symptoms were greater in COVID-positive than COVID-negative (38% *vs* 19%; *p=*0.001 and 23% *vs* 9.0 %; *p=*0.002, respectively). The proportion of patients requiring a medical consultation for persistent symptoms was similar between COVID-positive and COVID-negative (21% versus 15%, *p=*0.182).

**Table 3.** Factors associated with the presence of any long-term symptom during the phone survey in COVID-positive patients.

| Characteristics | No symptom<br>(n=195) | Any symptom<br>(n=223) | Bivariate analysis* |  | Multivariate<br>analysis*<sup>†</sup> |  |
| --- | --- | --- | --- | --- | --- | --- |
|  |  |  | OR<br>(95% CI) | P<br>value | aOR (95%<br>CI) | P<br>value |
| **Demographics** |  |  |  |  |  |  |
| Age in years | 41 (30-54) | 42 (33-54) | 1.01<br>(0.95-1.08) | 0.702 | 1.02<br>(0.94-1.10) | 0.682 |
| Female gender | 112 (57) | 149 (67) | 1.49<br>(1.00-2.22) | 0.049 | 1.67<br>(1.09-2.56) | 0.019 |
| Active smoking | 16 (8.2) | 30 (13) | 1.74<br>(0.92-3.30) | 0.090 | 1.72<br>(0.89-3.34) | 0.107 |
| **Comorbidities** |  |  |  |  |  |  |
| Overweight or obesity | 78 (40) | 112 (50) | 1.50<br>(1.02-2.21) | 0.041 | 1.67<br>(1.10-2.56) | 0.017 |
| Diabetes Mellitus | 12 (6.2) | 10 (4.5) | 0.72<br>(0.30-1.70) | 0.447 | 0.69<br>(0.28-1.72) | 0.426 |
| Asthma | 19 (9.7) | 24 (11) | 1.12<br>(0.59-2.11) | 0.732 | 1.11<br>(0.58-2.14) | 0.749 |
| Hypertension | 30 (15) | 35 (16) | 1.02<br>(0.60-1.74) | 0.930 | 0.96<br>(0.51-1.81) | 0.912 |
| Malignancy | 5 (2.6) | 10 (4.5) | 1.78<br>(0.60-5.31) | 0.298 | 1.63<br>(0.50-5.30) | 0.414 |
| Cardiovascular Disease | 5 (2.6) | 5 (2.2) | 0.87<br>(0.25-3.06) | 0.830 | 1.05<br>(0.27-4.09) | 0.948 |
| Chronic inflammatory<br>Disease | 5 (2.6) | 8 (3.6) | 1.41<br>(0.45-4.40) | 0.549 | 1.18<br>(0.35-3.94) | 0.790 |
| **Timing of phone**<br>**survey** |  |  |  |  |  |  |
| >3-5 months | 87 (45) | 103 (46) | 1.07<br>(0.72-1.57) | 0.747 | 1.06<br>(0.65-1.73) | 0.808 |
| >5-7 months | 47 (24) | 55 (25) | 1.03<br>(0.66-1.61) | 0.894 | 1.05<br>(0.61-1.80) | 0.856 |
| >7-10 months | 61 (31) | 65 (29) | 0.90<br>(0.59-1.37) | 0.635 | 1.00<br>(reference) | -- |

Data are number (%) of patients or median (interquartile range). Odds ratio for age indicates the risk of the presence of any symptom per 5-year age increase.
\*symptoms associated with the presence of any long-term symptom. <sup>†</sup>Analysis adjusted for age, gender, smoking, overweight/obesity, diabetes, asthma, hypertension, cancer, cardiovascular disease, chronic inflammatory disease, period of the phone survey.
Missing data: 1 BMI
OR: odds ratio; aOR: adjusted odds ratio; CI: confidence interval.

### Independent association between COVID-positive status and long-term symptoms

After adjusting for potential confounders, COVID-positive status was a predictor for reporting any symptom (adjusted odds ratio [aOR] 2.04, 95% CI 1.14-3.67; *p=*0.02), fatigue (aOR 2.14, 95% CI 1.04-4.41; *p=*0.04), dyspnea (aOR 2.81, 95% CI 1.10-7.16; *p=*0.03), smell or taste disorder (aOR 26.5, 95% CI 3.46-202; p<0.01) and memory impairment (aOR 5.71, 95% CI 1.53-21.3; *p=*0.01) at the time of the phone survey (Table 2). Of note, dyspnea associated with limited physical activity (NYHA ≥2), as well as other long-term symptoms were not associated with COVID status. The results were similar when restricting the analysis to COVID-negative and COVID-positive surveyed during third period (Appendix table 2).

### Predictors of long-term symptoms in COVID-positive

While 57% of women presented long-term symptoms, only 47% of men did (*p=*0.049). While 59% of overweight/obese patients presented long-term symptoms, only 49% of patients with healthy weight did (*p=*0.041). After adjusting for potential confounders, independent predictors of reporting any long-term symptoms were female gender (aOR 1.67, 95% CI 1.09-2.56; *p=*0.019) and overweight/obesity (aOR 1.67, 95% CI 1.10-2.56; *p=*0.017; Table 3). The same predictors were associated with long-term fatigue. Overweight/obesity was a predictor of dyspnea. Female gender was a predictor of smell or taste disorder. Active smoking was a predictor of memory impairment (Figure 3). Predictors of other long-term symptoms that were not associated with COVID status are presented in Appendix table 4.

**Figure 3:**
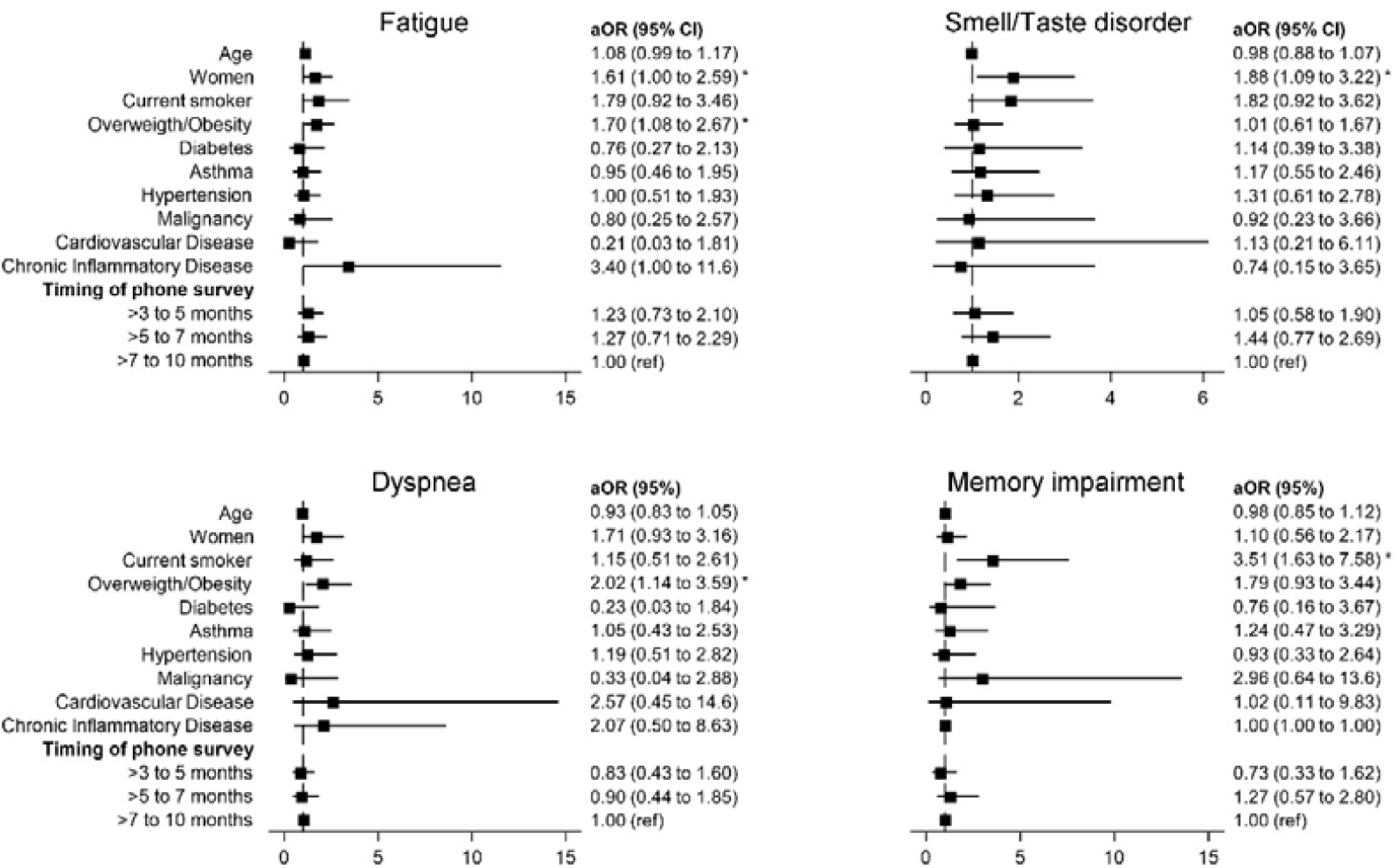
Factors associated with the presence of selected long-term symptoms (fatigue, smell or taste disorder, dyspnea and memory impairment) during the phone survey in COVID-positive patients. Selected symptoms are those associated with COVID status. Multivariable analysis adjusted for age, gender, smoking, overweight/obesity, diabetes, asthma, hypertension, cancer, cardiovascular disease, chronic inflammatory disease, period of the phone survey. Adjusted odds ratio for age indicates the risk of fatigue, smell/taste disorder, dyspnea or memory impairment per 5-year age increase. aOR: adjusted odds ratio; CI: confidence interval. *p<0.05

## DISCUSSION

In a large cohort of PCR-confirmed COVID-19 outpatients, we evaluated the prevalence of persistent symptoms 3 to 10 months after diagnosis. We compared COVID-positive to a cohort of patients with negative SARS-CoV-2 PCR with symptoms suggestive of COVID during the same period in the same study sites. In these populations consisting mostly of young and healthy participants, most working as HCW, we showed that 53% of patients with PCR-confirmed COVID-19 report persistent symptoms, while 37% with negative SARS-CoV-2 PCR do. Interestingly, only 4 out of the 14 surveyed persistent symptoms were independently associated with COVID status, i.e., fatigue, smell or taste disorder, dyspnea and memory impairment, irrespective of the timing of the survey.

The type and prevalence of long-lasting symptoms in COVID-positive are consistent with the existing literature. Two studies, evaluating more than 100 laboratory-confirmed COVID-19 outpatients, reported the prevalence of symptoms lasting more than six months after diagnosis: 33-46% participants with at least one symptom, 14-22% with fatigue, 8% with dyspnea, and 14-15% with change in sense of smell or taste (9,10). The differences between studies may be explained by different selection criteria, data collection methods, and definitions of symptoms.

Strikingly, in our study, the proportion of COVID-positive patients with long-term symptoms was similar between the three surveyed periods. It suggests that symptoms which are present three months after a COVID-19 diagnosis may last for several months. Our findings are supported by a couple of longitudinal studies, which analyzed the evolution of residual symptoms and showed a stable proportion after the acute phase up to seven months (17,18). In our study 23% of COVID-positive required at least one additional medical visit for COVID-19-related symptoms. These data show the burden on the ambulatory health-care facilities that may be related to the post-COVID-19 syndrome. In contrast with previous findings (19), we did not find a statistical difference in the proportion of patients requiring an additional medical consultation between COVID-positive and COVID-negative, which may be related to the timing of the survey, COVID-negative being surveyed at a later time point.

Our results shed more light on the association between SARS-CoV-2 infection and long-term symptoms. While some symptoms were more frequent in PCR-confirmed COVID-19 outpatients than in COVID-negative, the prevalence of other symptoms was not different between both groups. Specifically, the prevalence of headache and sleep disorders, two predominant persistent symptoms in COVID-negative patients, was similar between COVID-negative and COVID-positive. Both symptoms may originate from mixed etiologies including psychosocial factors and/or non-specific post-infectious consequences. The COVID-19 pandemic has greatly impacted our psychosocial behaviors due to confinement, social distancing, masks and changes in working conditions, especially for HCW. Some symptoms may be a consequence of these unprecedented changes (20).

One study evaluating the sequelae in outpatients six months after COVID-19 included a small control group of healthy volunteers and showed that 33% of COVID-positive outpatients and 5% of volunteers presented symptoms (9). However, the limited sample size (21 healthy volunteers) and normal health status prevents comparison with our results. Another report also showed that 26% of seropositive HCW present long-term symptoms eight months after mild COVID, while only 9% of seronegative do (21). Inclusion of patients irrespective of the presence of symptoms (no or mild prior symptoms) might explain the lower prevalence compared to our results.

Overweight and obesity are increasingly associated with poor outcomes in COVID-19 (22,23) and we found that it was an independent predictor for the presence of any long-term symptom, fatigue, and dyspnea. One study reported an association between body mass index and persistent symptoms after more than 28 days (24). The mechanisms responsible for the post-COVID-19 syndrome are not understood yet and may be multiple (25,26). Long-term symptoms may be due to higher initial organ damages related to a higher infectious burden or a dysregulated inflammatory response. This may be true in people with overweight and obesity (27), since the adipose tissue has a high expression of angiotensin-converting enzyme 2 receptors and secretes pro-inflammatory cytokines.

We identified female gender as a predictor for the post-COVID-19 syndrome, in particular for persistent fatigue and smell or taste disorder. Although the results are inconsistent, some studies have described an association between female gender and long-COVID (24), persistent fatigue (7,8,28), post-exertional polypnea (28), and anxiety or depression (8). There may be a difference in the immune response according to the gender, as illustrated by the higher representation of women in autoimmune diseases (29), that may explain divergent findings with COVID-19.

Our study has some limitations. First, most symptoms are subjective and prone to observer bias. Symptoms may also come from an intercurrent condition at the time of COVID-19 diagnosis and not always represent sequelae of SARS-CoV-2 infection. However, inclusion of a symptomatic COVID-negative group supports the association between some persistent symptoms and SARS-CoV-2 infection. Second, inclusion criteria were slightly different between COVID-positive and COVID-negative. COVID-positive presented with any symptom suggestive of COVID-19 (cough, dyspnea, sore throat, history of fever, myalgia, or smell or taste disorders), while the inclusion criteria of COVID-negative patients were the presence of cough, dyspnea or history of fever. However, we do not think that the symptoms difference at first consultation modified our results. Indeed, among COVID-positive patients, 361/418 (86%) presented with cough, dyspnea or history of fever, which were the inclusion criteria of the COVID-negative group. Another limitation is related to the timing of the survey, which was performed later in COVID-negative. This difference could lead to an underestimation of the prevalence of persisting symptoms in COVID-negative. However, we added a subgroup analysis restricted to COVID-positive and COVID-negative surveyed during the same period (the third one) and found the same results than in the whole cohort.

We also observed a difference in the characteristics of COVID-positive surveyed during the first period compared to COVID-positive surveyed during the other two periods. This difference is the result of chance as patients as clinicians called patients randomly. However, we adjusted the multivariate analyses for these characteristics and for the timing of the survey. Our study suffers from a selection bias due to SARS-CoV-2 test criteria (HCW or presence of a risk factor of adverse outcome) at the time of the study, which may prevent generalization of our findings to a broader population. We added a subgroup analysis restricted to HCW and found the same results than in the complete population. This information suggests that our results are generalizable to HCW. Furthermore, we cannot formally exclude that some COVID-negative had an undiagnosed SARS-CoV-2 infection due to a false negative PCR result. However, we used a validated SARS-CoV-2 RT-PCR test on a nasopharyngeal swab performed by a dedicated trained medical team to minimize technical and sample collection bias (12,30).

In conclusion, our study shows that more than half of outpatients with mild-to-moderate COVID-19 report long-term symptoms 3 to 10 months after diagnosis and that 21% seek medical care for this reason. These data suggest that the post-COVID syndrome places a significant burden on society and especially healthcare systems. There is an urgent need to inform physicians and political authorities about the natural long-term course of COVID-19 in order to plan an appropriate and dedicated management of those with disabling persistent symptoms.

## Data Availability

The data that support the findings of this study are available from the corresponding author, FD, upon reasonable request.

## Acknowledgements

We thank all the patients who accepted to participate and make this study possible. We thank all healthcare workers of the triage center and of the emergency department of the University Hospital of Lausanne, who supported the study and managed COVID-19 suspected patients. We thank Nadia Cattaneo and Martin Delaloye who participated to patients’ follow-up. We thank Tanguy Espejo and Luca Bosso who included patients in the COVID-negative group. Members of the RegCOVID group (by alphabetical order): Pierre-Yves Bochud (Committee President), Florian Desgranges, Paraskevas Filippidis, David Haefliger, Eleftheria Evdokia Kampouri, Oriol Manuel (Committee Member), Aline Munting, Jean-Luc Pagani (Committee Member), Matthaios Papadimitriou-Olivgeris (Registry Coordinator and Committee Member), Jean Regina, Laurence Rochat Stettler, Véronique Suttels, Eliana Tadini, Jonathan Tschopp, Mathias Van Singer, Benjamin Viala, Peter Vollenweider (Committee member).

## Authors’ contributions

FD, MPO, NBB: study conception, study design, study performance, study management, patients’ follow-up, data analysis, data interpretation and manuscript writing.

ET, AM: patients follow-up, data interpretation and manuscript writing.

JR, PF, BV, NS, DH, EK, MVS, JT, LRS: patients follow-up, data interpretation and critical review of the manuscript.

BG: study design, data interpretation and critical review of the manuscript.

OH, YMC, AG, PNC, BG: data interpretation and critical review of the manuscript.

EK: data analysis, interpretation, and critical review of the manuscript.

TB, SS: patients’ inclusion, and critical review of the manuscript.

OO: microbiological data interpretation and critical review of the manuscript

MPO had full access to all the data in the study and takes responsibility for the integrity of the data and the accuracy of the data analysis.

### APPENDICES

**Appendix table 1.**
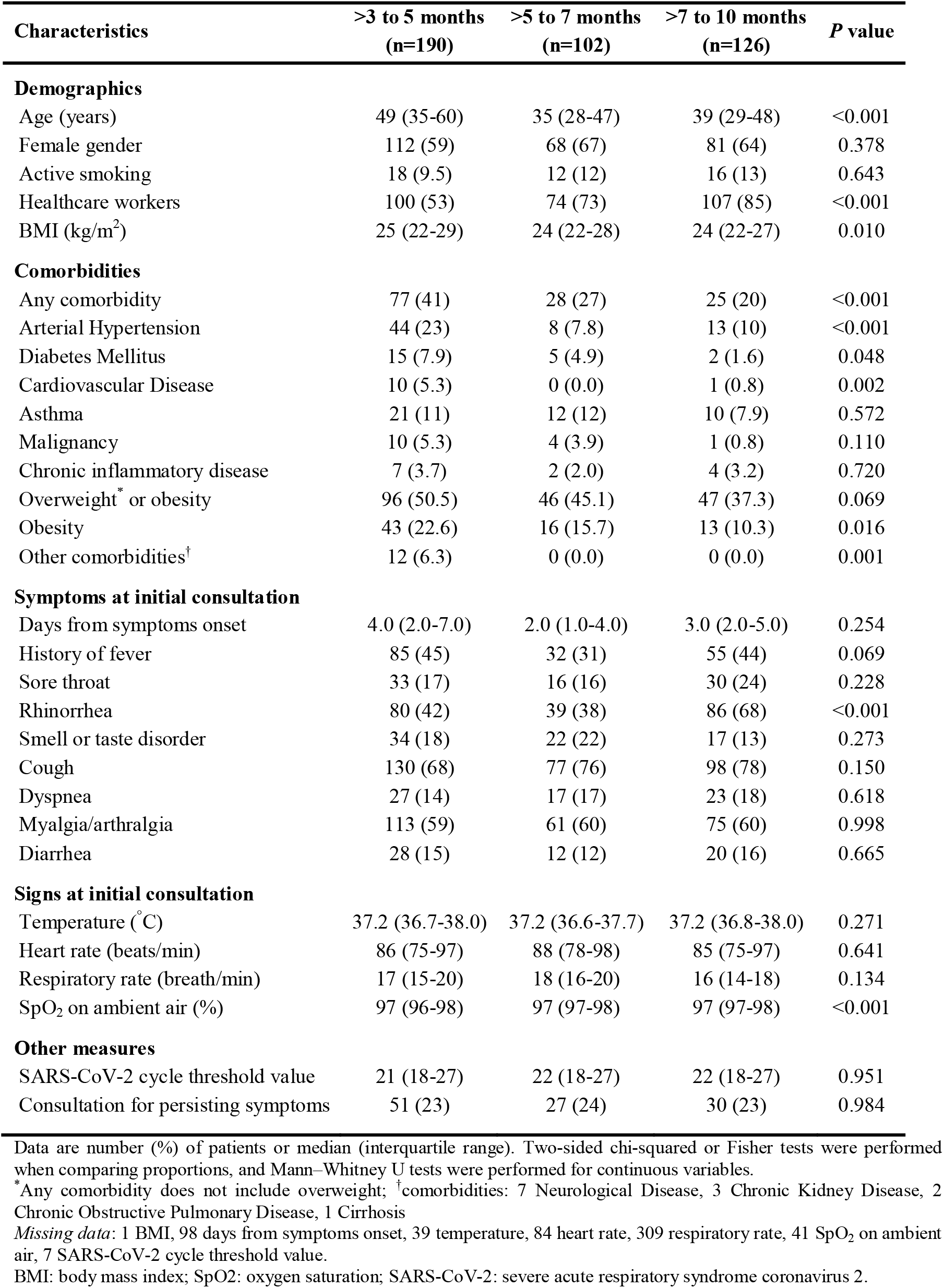
Characteristics of COVID-positive patients according to the period of the phone interview (time between SARS-CoV-2 diagnosis and phone interview).

**Appendix table 2.**
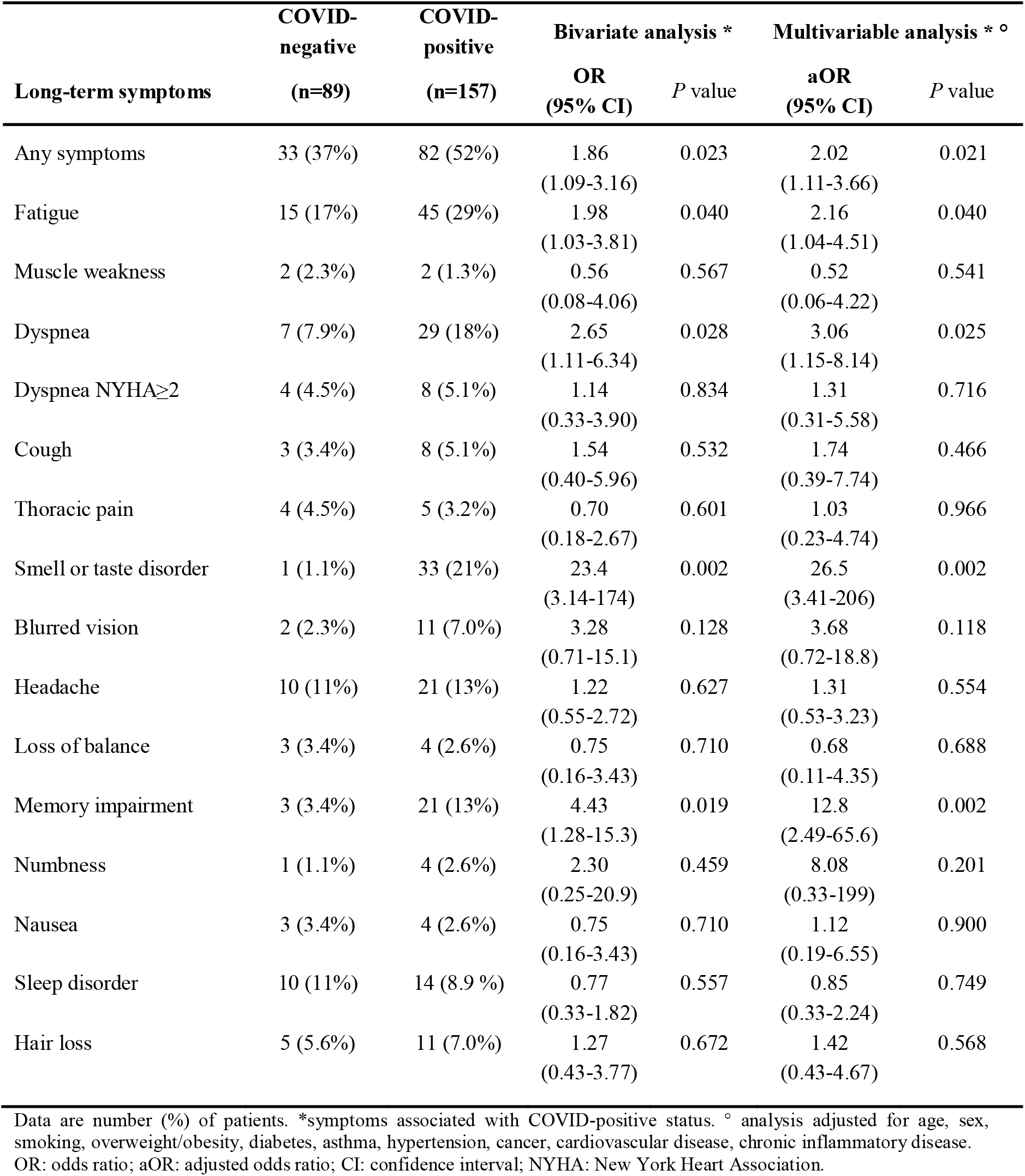
Association between long-term symptoms (any and individualized symptoms) assessed at the time of phone survey and COVID-positive status among patients surveyed during the third period.

**Appendix table 3A:** Characteristics of COVID-positive patients and proportion of COVID-positive patients with long-term symptoms (any and individualized symptoms) assessed at the time of phone survey according to health care worker (HCW) status. Data are number (%) of patients or median (interquartile range). Two-sided chi-squared or Fisher tests were performed when comparing proportions, and Mann–Whitney U tests were performed for continuous variables.

| Characteristics | Not HCW (N=137) | HCW (N=281) | P value |
| --- | --- | --- | --- |
| **Demographics** |  |  |  |
| Age in years | 51 (38-64) | 38 (30-48) | <0.001 |
| Female gender | 66 (48) | 195 (69) | <0.001 |
| Active smoking | 14 (10) | 32 (11) | 0.720 |
| BMI (kg/m<sup>2</sup>) | 26 (23-29) | 24 (21-28) | <0.001 |
| **Comorbidities** |  |  |  |
| Any comorbidity | 77 (56) | 53 (19) | <0.001 |
| Arterial Hypertension | 40 (29) | 25 (8.9) | <0.001 |
| Diabetes Mellitus | 15 (11) | 7 (2.5) | <0.001 |
| Cardiovascular Disease | 7 (5.1) | 3 (1.1) | 0.017 |
| Asthma | 22 (16) | 21 (7.5) | 0.007 |
| Malignancy (hematological/solid organ) | 11 (8.0) | 4 (1.4) | 0.001 |
| Chronic inflammatory disease | 8 (5.8) | 5 (1.8) | 0.032 |
| Overweight* or obesity | 80 (58) | 109 (39) | <0.001 |
| Obesity | 31 (23) | 41 (15) | 0.041 |
| Other comorbidities† | 9 (6.6) | 3 (1.1) | 0.002 |
| **Long-term symptoms** |  |  |  |
| Any symptoms | 76 (56) | 147 (52) | 0.543 |
| Fatigue | 45 (33) | 87 (31) | 0.737 |
| Muscle weakness | 2 (1.5) | 6 (2.1) | 0.636 |
| Dyspnea | 22 (16) | 44 (16) | 0.916 |
| Dyspnea NYHA≥2 | 11 (8.0) | 19 (6.8) | 0.637 |
| Cough | 11 (8.0) | 14 (5.0) | 0.271 |
| Thoracic pain | 9 (6.6) | 12 (4.3) | 0.313 |
| Smell or taste disorder | 25 (18) | 68 (24) | 0.170 |
| Blurred vision | 7 (5.1) | 13 (4.6) | 0.288 |
| Headache | 18 (13) | 32 (11) | 0.605 |
| Loss of balance | 7 (5.1) | 5 (1.8) | 0.066 |
| Memory impairment | 16 (12) | 32 (11) | 0.930 |
| Numbness | 7 (5.1) | 7 (2.5) | 0.163 |
| Nausea | 7 (5.1) | 4 (1.4) | 0.045 |
| Sleep disorder | 15 (11) | 26 (9.3) | 0.584 |
| Hair loss | 14 (10) | 29 (10) | 0.974 |

\*Any comorbidity does not include overweight; †Other comorbidities: 7 Neurological Disease, 3 Chronic Kidney Disease, 2 Chronic Obstructive Pulmonary Disease, 1 Cirrhosis.
HCW: health care worker.

**Appendix table 3B.**
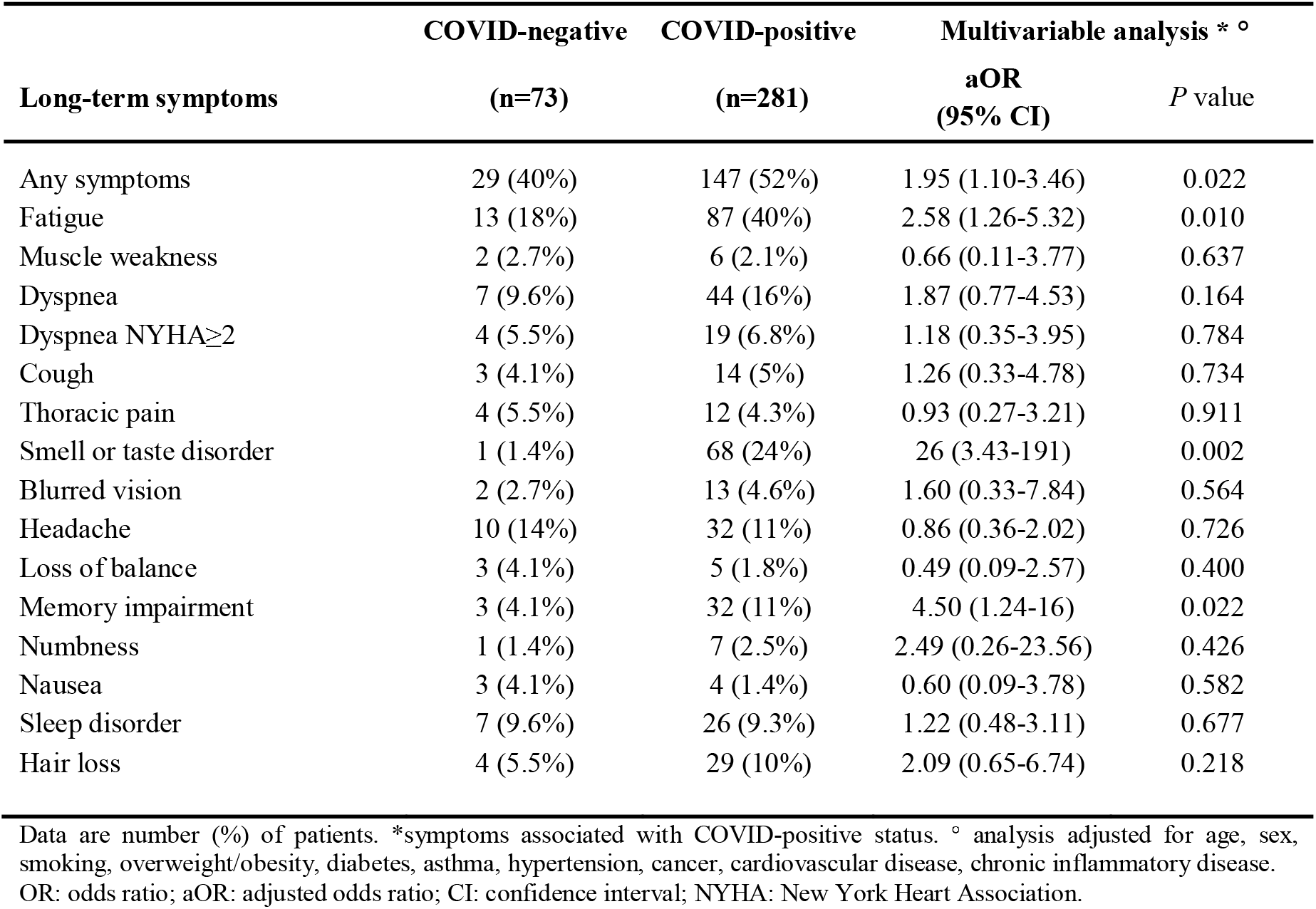
Association between long-term symptoms (any and individualized symptoms) assessed at the time of phone survey and COVID-positive status among health care workers.

**Appendix Table 4.**
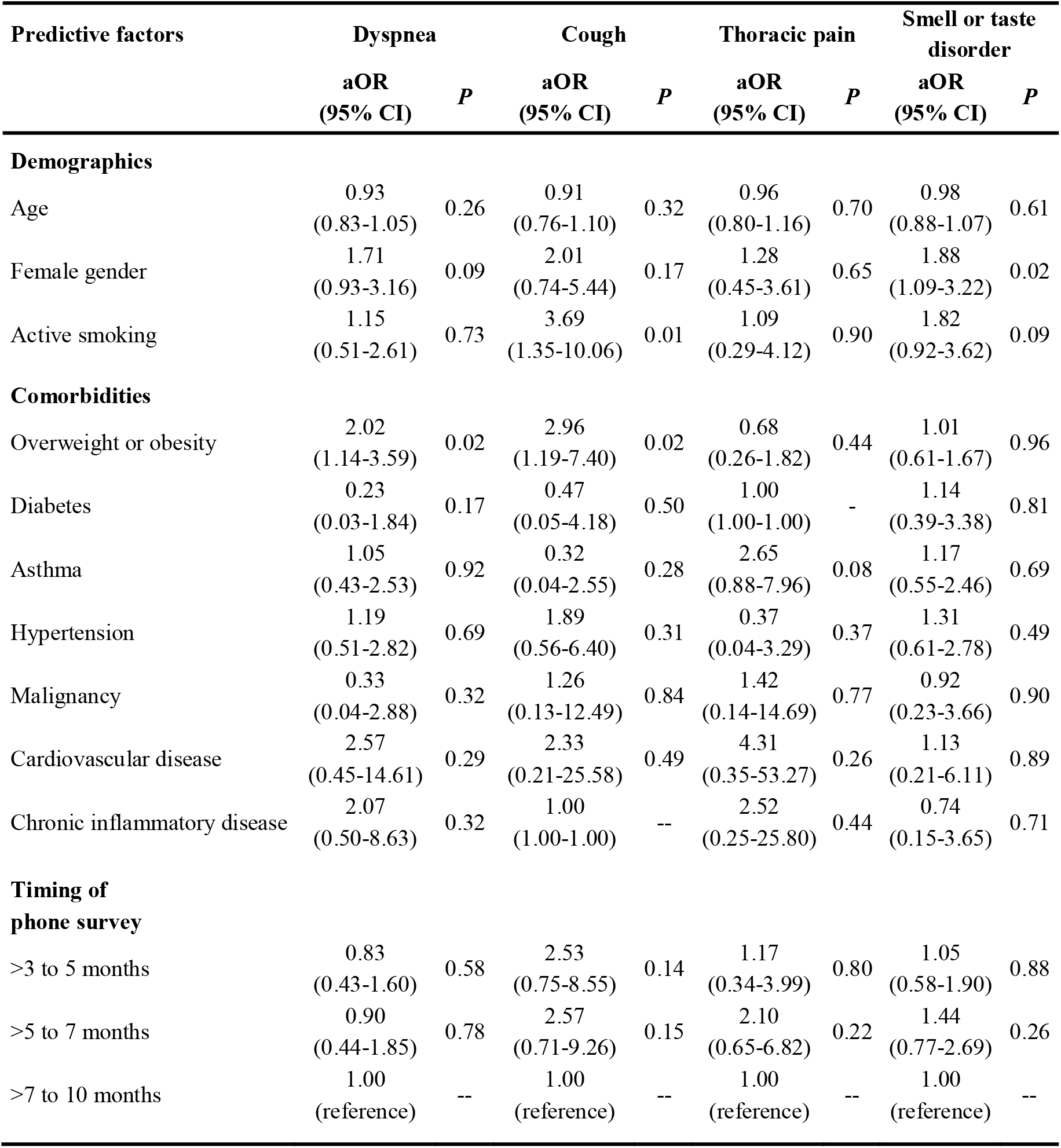
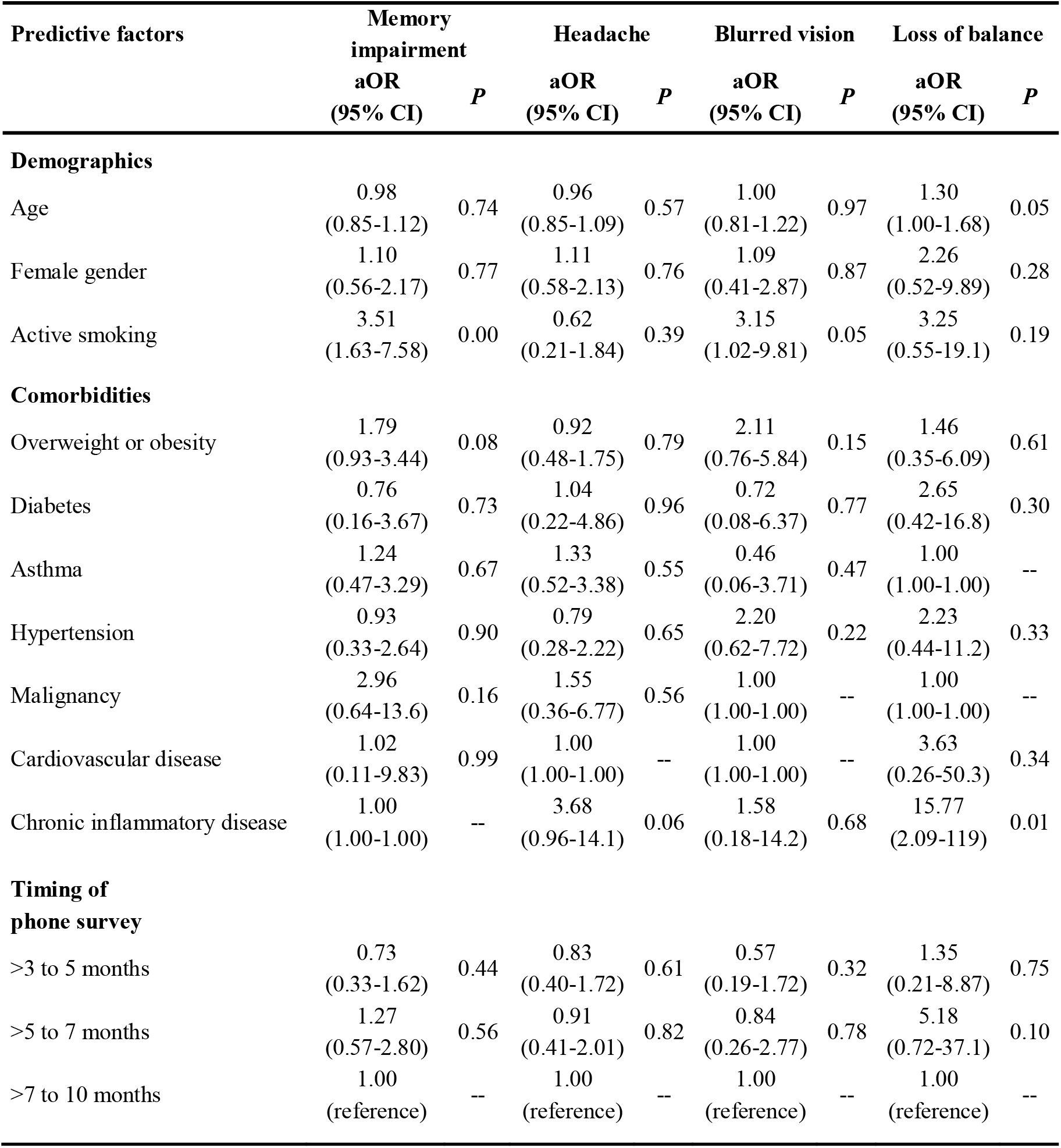
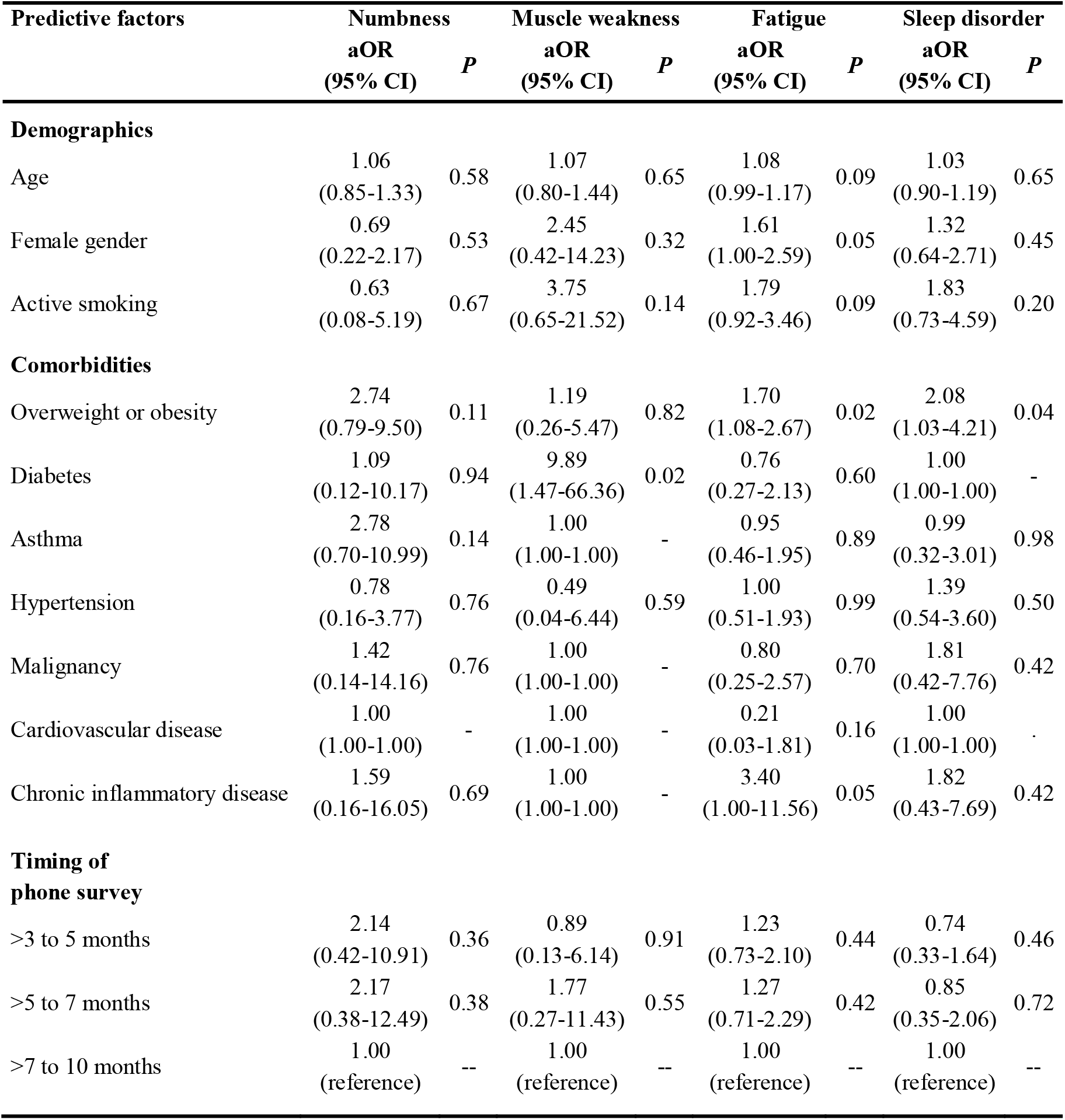
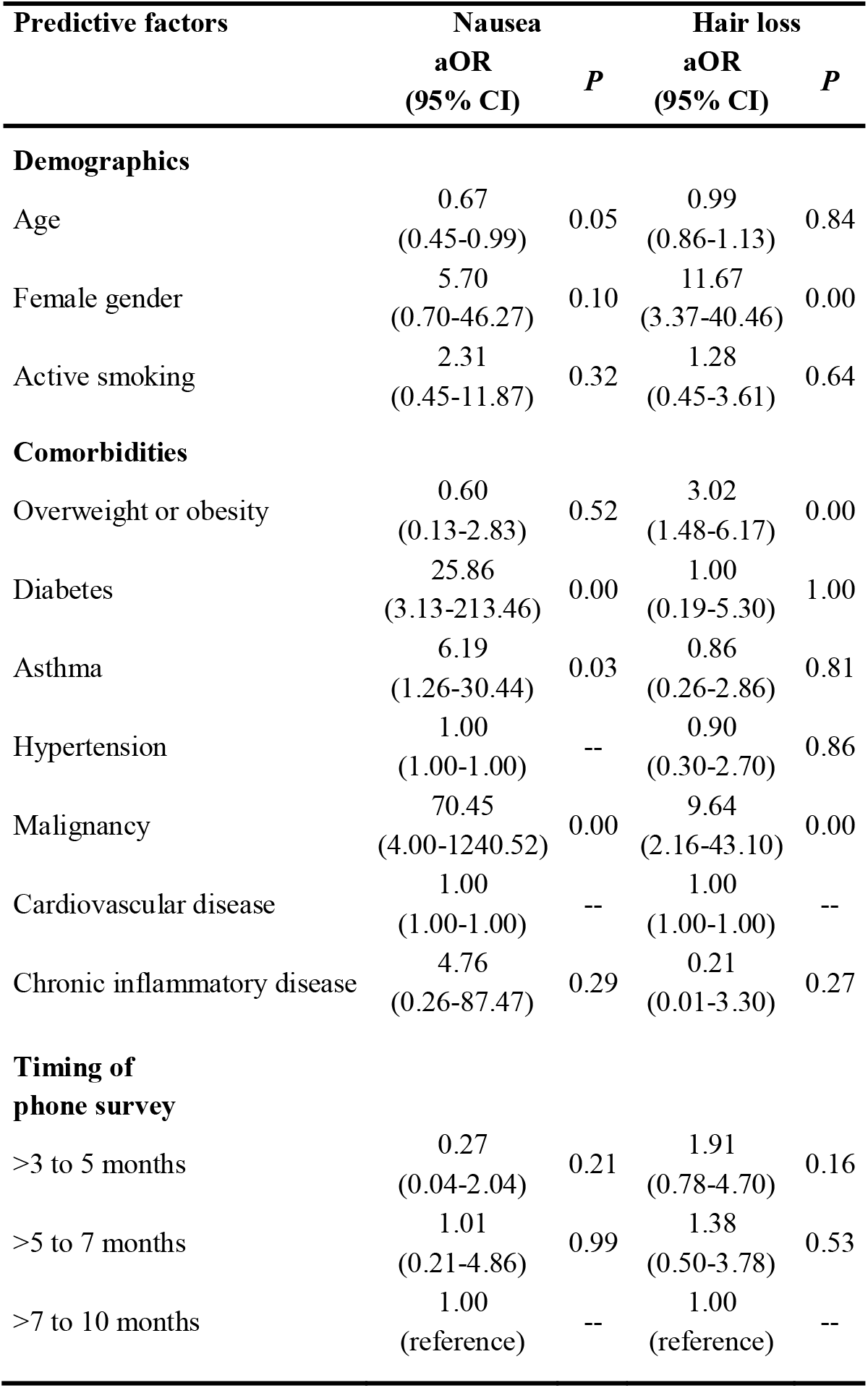
Factors associated with the presence of long-term symptoms during the phone survey in COVID-positive patients. Analysis adjusted for age, gender, smoking, overweight/obesity, diabetes, asthma, hypertension, cancer, cardiovascular disease, chronic inflammatory disease, period of the phone survey. *Missing data*: 1 BMI. Adjusted odds ratio for age indicates the risk of the presence of any symptom or individualized symptoms per 5-year age increase. aOR: adjusted odds ratio; CI: confidence interval.

| Predictive factors | Dyspnea |  | Cough |  | Thoracic pain |  | Smell or taste disorder |  |
| --- | --- | --- | --- | --- | --- | --- | --- | --- |
|  | aOR<br>(95% CI) | P | aOR<br>(95% CI) | P | aOR<br>(95% CI) | P | aOR<br>(95% CI) | P |
| **Demographics** |  |  |  |  |  |  |  |  |
| Age | 0.93<br>(0.83-1.05) | 0.26 | 0.91<br>(0.76-1.10) | 0.32 | 0.96<br>(0.80-1.16) | 0.70 | 0.98<br>(0.88-1.07) | 0.61 |
| Female gender | 1.71<br>(0.93-3.16) | 0.09 | 2.01<br>(0.74-5.44) | 0.17 | 1.28<br>(0.45-3.61) | 0.65 | 1.88<br>(1.09-3.22) | 0.02 |
| Active smoking | 1.15<br>(0.51-2.61) | 0.73 | 3.69<br>(1.35-10.06) | 0.01 | 1.09<br>(0.29-4.12) | 0.90 | 1.82<br>(0.92-3.62) | 0.09 |
| **Comorbidities** |  |  |  |  |  |  |  |  |
| Overweight or obesity | 2.02<br>(1.14-3.59) | 0.02 | 2.96<br>(1.19-7.40) | 0.02 | 0.68<br>(0.26-1.82) | 0.44 | 1.01<br>(0.61-1.67) | 0.96 |
| Diabetes | 0.23<br>(0.03-1.84) | 0.17 | 0.47<br>(0.05-4.18) | 0.50 | 1.00<br>(1.00-1.00) | - | 1.14<br>(0.39-3.38) | 0.81 |
| Asthma | 1.05<br>(0.43-2.53) | 0.92 | 0.32<br>(0.04-2.55) | 0.28 | 2.65<br>(0.88-7.96) | 0.08 | 1.17<br>(0.55-2.46) | 0.69 |
| Hypertension | 1.19<br>(0.51-2.82) | 0.69 | 1.89<br>(0.56-6.40) | 0.31 | 0.37<br>(0.04-3.29) | 0.37 | 1.31<br>(0.61-2.78) | 0.49 |
| Malignancy | 0.33<br>(0.04-2.88) | 0.32 | 1.26<br>(0.13-12.49) | 0.84 | 1.42<br>(0.14-14.69) | 0.77 | 0.92<br>(0.23-3.66) | 0.90 |
| Cardiovascular disease | 2.57<br>(0.45-14.61) | 0.29 | 2.33<br>(0.21-25.58) | 0.49 | 4.31<br>(0.35-53.27) | 0.26 | 1.13<br>(0.21-6.11) | 0.89 |
| Chronic inflammatory disease | 2.07<br>(0.50-8.63) | 0.32 | 1.00<br>(1.00-1.00) | -- | 2.52<br>(0.25-25.80) | 0.44 | 0.74<br>(0.15-3.65) | 0.71 |
| **Timing of phone survey** |  |  |  |  |  |  |  |  |
| >3 to 5 months | 0.83<br>(0.43-1.60) | 0.58 | 2.53<br>(0.75-8.55) | 0.14 | 1.17<br>(0.34-3.99) | 0.80 | 1.05<br>(0.58-1.90) | 0.88 |
| >5 to 7 months | 0.90<br>(0.44-1.85) | 0.78 | 2.57<br>(0.71-9.26) | 0.15 | 2.10<br>(0.65-6.82) | 0.22 | 1.44<br>(0.77-2.69) | 0.26 |
| >7 to 10 months | 1.00<br>(reference) | -- | 1.00<br>(reference) | -- | 1.00<br>(reference) | -- | 1.00<br>(reference) | -- |

| Predictive factors | Memory impairment |  | Headache |  | Blurred vision |  | Loss of balance |  |
| --- | --- | --- | --- | --- | --- | --- | --- | --- |
|  | aOR<br>(95% CI) | P | aOR<br>(95% CI) | P | aOR<br>(95% CI) | P | aOR<br>(95% CI) | P |
| **Demographics** |  |  |  |  |  |  |  |  |
| Age | 0.98<br>(0.85-1.12) | 0.74 | 0.96<br>(0.85-1.09) | 0.57 | 1.00<br>(0.81-1.22) | 0.97 | 1.30<br>(1.00-1.68) | 0.05 |
| Female gender | 1.10<br>(0.56-2.17) | 0.77 | 1.11<br>(0.58-2.13) | 0.76 | 1.09<br>(0.41-2.87) | 0.87 | 2.26<br>(0.52-9.89) | 0.28 |
| Active smoking | 3.51<br>(1.63-7.58) | 0.00 | 0.62<br>(0.21-1.84) | 0.39 | 3.15<br>(1.02-9.81) | 0.05 | 3.25<br>(0.55-19.1) | 0.19 |
| **Comorbidities** |  |  |  |  |  |  |  |  |
| Overweight or obesity | 1.79<br>(0.93-3.44) | 0.08 | 0.92<br>(0.48-1.75) | 0.79 | 2.11<br>(0.76-5.84) | 0.15 | 1.46<br>(0.35-6.09) | 0.61 |
| Diabetes | 0.76<br>(0.16-3.67) | 0.73 | 1.04<br>(0.22-4.86) | 0.96 | 0.72<br>(0.08-6.37) | 0.77 | 2.65<br>(0.42-16.8) | 0.30 |
| Asthma | 1.24<br>(0.47-3.29) | 0.67 | 1.33<br>(0.52-3.38) | 0.55 | 0.46<br>(0.06-3.71) | 0.47 | 1.00<br>(1.00-1.00) | -- |
| Hypertension | 0.93<br>(0.33-2.64) | 0.90 | 0.79<br>(0.28-2.22) | 0.65 | 2.20<br>(0.62-7.72) | 0.22 | 2.23<br>(0.44-11.2) | 0.33 |
| Malignancy | 2.96<br>(0.64-13.6) | 0.16 | 1.55<br>(0.36-6.77) | 0.56 | 1.00<br>(1.00-1.00) | -- | 1.00<br>(1.00-1.00) | -- |
| Cardiovascular disease | 1.02<br>(0.11-9.83) | 0.99 | 1.00<br>(1.00-1.00) | -- | 1.00<br>(1.00-1.00) | -- | 3.63<br>(0.26-50.3) | 0.34 |
| Chronic inflammatory disease | 1.00<br>(1.00-1.00) | -- | 3.68<br>(0.96-14.1) | 0.06 | 1.58<br>(0.18-14.2) | 0.68 | 15.77<br>(2.09-119) | 0.01 |
| **Timing of phone survey** |  |  |  |  |  |  |  |  |
| >3 to 5 months | 0.73<br>(0.33-1.62) | 0.44 | 0.83<br>(0.40-1.72) | 0.61 | 0.57<br>(0.19-1.72) | 0.32 | 1.35<br>(0.21-8.87) | 0.75 |
| >5 to 7 months | 1.27<br>(0.57-2.80) | 0.56 | 0.91<br>(0.41-2.01) | 0.82 | 0.84<br>(0.26-2.77) | 0.78 | 5.18<br>(0.72-37.1) | 0.10 |
| >7 to 10 months | 1.00<br>(reference) | -- | 1.00<br>(reference) | -- | 1.00<br>(reference) | -- | 1.00<br>(reference) | -- |

| Predictive factors | Numbness |  | Muscle weakness |  | Fatigue |  | Sleep disorder |  |
| --- | --- | --- | --- | --- | --- | --- | --- | --- |
|  | aOR<br>(95% CI) | P | aOR<br>(95% CI) | P | aOR<br>(95% CI) | P | aOR<br>(95% CI) | P |
| **Demographics** |  |  |  |  |  |  |  |  |
| Age | 1.06<br>(0.85-1.33) | 0.58 | 1.07<br>(0.80-1.44) | 0.65 | 1.08<br>(0.99-1.17) | 0.09 | 1.03<br>(0.90-1.19) | 0.65 |
| Female gender | 0.69<br>(0.22-2.17) | 0.53 | 2.45<br>(0.42-14.23) | 0.32 | 1.61<br>(1.00-2.59) | 0.05 | 1.32<br>(0.64-2.71) | 0.45 |
| Active smoking | 0.63<br>(0.08-5.19) | 0.67 | 3.75<br>(0.65-21.52) | 0.14 | 1.79<br>(0.92-3.46) | 0.09 | 1.83<br>(0.73-4.59) | 0.20 |
| **Comorbidities** |  |  |  |  |  |  |  |  |
| Overweight or obesity | 2.74<br>(0.79-9.50) | 0.11 | 1.19<br>(0.26-5.47) | 0.82 | 1.70<br>(1.08-2.67) | 0.02 | 2.08<br>(1.03-4.21) | 0.04 |
| Diabetes | 1.09<br>(0.12-10.17) | 0.94 | 9.89<br>(1.47-66.36) | 0.02 | 0.76<br>(0.27-2.13) | 0.60 | 1.00<br>(1.00-1.00) | - |
| Asthma | 2.78<br>(0.70-10.99) | 0.14 | 1.00<br>(1.00-1.00) | - | 0.95<br>(0.46-1.95) | 0.89 | 0.99<br>(0.32-3.01) | 0.98 |
| Hypertension | 0.78<br>(0.16-3.77) | 0.76 | 0.49<br>(0.04-6.44) | 0.59 | 1.00<br>(0.51-1.93) | 0.99 | 1.39<br>(0.54-3.60) | 0.50 |
| Malignancy | 1.42<br>(0.14-14.16) | 0.76 | 1.00<br>(1.00-1.00) | - | 0.80<br>(0.25-2.57) | 0.70 | 1.81<br>(0.42-7.76) | 0.42 |
| Cardiovascular disease | 1.00<br>(1.00-1.00) | - | 1.00<br>(1.00-1.00) | - | 0.21<br>(0.03-1.81) | 0.16 | 1.00<br>(1.00-1.00) | - |
| Chronic inflammatory disease | 1.59<br>(0.16-16.05) | 0.69 | 1.00<br>(1.00-1.00) | - | 3.40<br>(1.00-11.56) | 0.05 | 1.82<br>(0.43-7.69) | 0.42 |
| **Timing of phone survey** |  |  |  |  |  |  |  |  |
| >3 to 5 months | 2.14<br>(0.42-10.91) | 0.36 | 0.89<br>(0.13-6.14) | 0.91 | 1.23<br>(0.73-2.10) | 0.44 | 0.74<br>(0.33-1.64) | 0.46 |
| >5 to 7 months | 2.17<br>(0.38-12.49) | 0.38 | 1.77<br>(0.27-11.43) | 0.55 | 1.27<br>(0.71-2.29) | 0.42 | 0.85<br>(0.35-2.06) | 0.72 |
| >7 to 10 months | 1.00<br>(reference) | -- | 1.00<br>(reference) | -- | 1.00<br>(reference) | -- | 1.00<br>(reference) | -- |

| Predictive factors | Nausea |  | Hair loss |  |
| --- | --- | --- | --- | --- |
|  | aOR<br>(95% CI) | *P* | aOR<br>(95% CI) | *P* |
| **Demographics** |  |  |  |  |
| Age | 0.67<br>(0.45-0.99) | 0.05 | 0.99<br>(0.86-1.13) | 0.84 |
| Female gender | 5.70<br>(0.70-46.27) | 0.10 | 11.67<br>(3.37-40.46) | 0.00 |
| Active smoking | 2.31<br>(0.45-11.87) | 0.32 | 1.28<br>(0.45-3.61) | 0.64 |
| **Comorbidities** |  |  |  |  |
| Overweight or obesity | 0.60<br>(0.13-2.83) | 0.52 | 3.02<br>(1.48-6.17) | 0.00 |
| Diabetes | 25.86<br>(3.13-213.46) | 0.00 | 1.00<br>(0.19-5.30) | 1.00 |
| Asthma | 6.19<br>(1.26-30.44) | 0.03 | 0.86<br>(0.26-2.86) | 0.81 |
| Hypertension | 1.00<br>(1.00-1.00) | -- | 0.90<br>(0.30-2.70) | 0.86 |
| Malignancy | 70.45<br>(4.00-1240.52) | 0.00 | 9.64<br>(2.16-43.10) | 0.00 |
| Cardiovascular disease | 1.00<br>(1.00-1.00) | -- | 1.00<br>(1.00-1.00) | -- |
| Chronic inflammatory disease | 4.76<br>(0.26-87.47) | 0.29 | 0.21<br>(0.01-3.30) | 0.27 |
| **Timing of**<br>**phone survey** |  |  |  |  |
| >3 to 5 months | 0.27<br>(0.04-2.04) | 0.21 | 1.91<br>(0.78-4.70) | 0.16 |
| >5 to 7 months | 1.01<br>(0.21-4.86) | 0.99 | 1.38<br>(0.50-3.78) | 0.53 |
| >7 to 10 months | 1.00<br>(reference) | -- | 1.00<br>(reference) | -- |

